# Systematic review and meta-analysis to estimate the burden of fatal and non-fatal overdose among people who inject drugs

**DOI:** 10.1101/2022.02.18.22271192

**Authors:** Jalissa Shealey, Eric W. Hall, Therese D. Pigott, Heather Bradley

## Abstract

**Background:** People who inject drugs (PWID) have high overdose risk. To assess the burden of drug overdose among PWID in light of opioid epidemic-associated increases in injection drug use (IDU), we estimated rates of non-fatal and fatal overdose among PWID living in Organization for Economic Cooperation and Development (OECD) countries using data from 2010 or later.

**Methods:** PubMed, Psych Info, and Embase databases were systematically searched to identify peer-reviewed studies reporting prevalence or rates of recent (past 12 months) fatal or non-fatal overdose events among PWID in OECD countries. Data were extracted and meta-analyzed using random effects models to produce pooled non-fatal and fatal overdose rates.

**Results:** 57 of 13,307 identified reports were included in the review, with 33/57 studies contributing unique data and included in the meta-analysis. Other (24/57) studies presented overlapping data to those included in meta-analysis. The rates of non-fatal and fatal overdose among PWID in OECD countries were 24.74 per 100 person years (PY) (95% CI: 19.86 – 30.83; n=28; I^2^=98.5%) and 0.61 per 100 PY (95% CI: 0.32 – 1.16; n=8; I^2^=93.4%), respectively. The rate of non-fatal overdose was 27.79 in North American countries, 25.71 in Canada, 28.59 in the U.S., and 21.44 in Australia.

**Conclusion:** These findings suggest there is a fatal overdose for every 40 non-fatal overdose events among PWID in OECD countries. The magnitude of overdose burden estimated here underscores the need for expansion of overdose prevention and treatment programs and serves as a baseline estimate for monitoring success of such programs.

## Introduction

People who inject drugs (PWID) have high risk for overdose, leading to adverse health outcomes including preventable death. It is estimated that, globally, 3.2 million PWID experience at least one non-fatal overdose over the course of a year (Colledge et al., 2019). Compared to people who use substances via other consumption routes, PWID are more likely to experience both non-fatal and fatal overdose due to rapid onset of effects (Caudarella et al., 2016; Mars et al., 2018). Physical consequences of non-fatal overdose can be severe (e.g., cognitive impairment, peripheral neuropathy, paralysis of limbs) and result in lifelong morbidity (Park et al., 2020; Warner-Smith et al., 2002). Consequently, healthcare utilization costs of overdose are substantial. In 2017, the economic burden of overdose and opioid use disorder (OUD) in the U.S. was estimated at $1.02 trillion, with more than 85% of costs attributable to the value of reduced quality of life and loss of life (Florence et al., 2021). Most concerning, non-fatal overdose events may be followed by subsequent fatal overdose (Caudarella et al., 2016).

Injection drug use (IDU) has likely increased during the past decade due to the trajectory of the U.S. opioid epidemic. While the first wave (1990-2009) of the opioid epidemic involved primarily prescription opioids, the second (2010-2012) and third (2013-) waves are characterized to a large extent by use of primarily injected substances (e.g., heroin, synthetic opioids including fentanyl) (CDC, 2021d). Increasing IDU is difficult to measure directly but is indicated by observed increases in infectious diseases primarily affecting PWID, such as acute hepatitis C virus (CDC, 2021a; Zibbell et al., 2018), infectious endocarditis, and skin and soft tissue infections (Dahlman et al., 2017; Weir et al., 2019). Increases in HIV transmissions among PWID have also been observed in the context of large HIV outbreaks (Alpren et al., 2020; Conrad et al., 2015; Peters et al., 2016). There is an urgent need to understand the burden of non-fatal and fatal overdose in the context of increased IDU.

Monitoring the burden of non-fatal and fatal overdose among PWID is important because overdose events can be prevented with evidence-based interventions. Harm reduction interventions such as syringe services programs (SSPs) and naloxone distribution, as well as medication-assisted treatment (MAT) when appropriate, reduce risk for overdose and other adverse health outcomes among PWID (Adams et al., 2019; Buresh et al., 2020; Ma et al., 2019). Overdose burden estimates can inform resource allocation for prevention programs and are needed for monitoring prevention effectiveness over time.

Given the relative sparsity of data on overdose among PWID, meta-analyses are important tools to produce summary estimates of non-fatal and fatal overdose rates. Here, we use the same meta-analytic methods to estimate non-fatal and fatal overdose rates, facilitating estimation of a fatal to non-fatal overdose ratio that can be used to extrapolate the number of fatal overdoses to a corresponding number of non-fatal events. This meta-analysis also provides updates to several previously conducted, which include data prior to the second and third opioid epidemic waves (pre-2010). For example, Colledge et al. (2019) conducted a global systematic review and meta-analysis of non-fatal overdose among PWID, including studies published during 2002 – 2018 (Colledge et al., 2019). Other related meta-analyses focused more broadly on all-cause mortality among PWID (Mathers et al., 2013) or included people with opioid use disorder regardless of primary route of administration (Bahji et al., 2020; Colledge et al., 2019; Larney et al., 2020; Mathers et al., 2013).

We used data collected in 2010 or later from PWID in Organization for Economic Cooperation and Development (OECD) countries, which have been relatively similarly affected by the opioid epidemic, to estimate overall and country-specific rates of non-fatal and fatal overdose in OECD countries.

## Methods

### 2.1 Search Strategy

This systematic review and meta-analysis were conducted in accordance with methods outlined in the 2020 Preferred Reporting Items for Systematic Reviews and Meta-Analyses (PRISMA) statement guidelines (Page et al., 2021). PubMed, Psych Info, and Embase databases were systematically searched to identify peer-reviewed studies on overdose among PWID published from January 1, 2010 to August 28, 2020. Search terms contained a term related to drug injection (e.g., “PWID”, “IDU”, “injection”) and a term for overdose or death (e.g., “mortality”, “fatal”). Complete details of the search terms can be found in Appendix 1.

We imported all search results into Covidence, systematic review management software, and removed duplicate citations. Titles and abstracts were each screened by one team member (J.Y.S., H.B., E.H., and M.G.). Next, the full text manuscripts were independently screened for eligibility by pairs of four researchers (J.Y.S., H.B., E.H., and M.G.), and conflicts were resolved collaboratively or by a third reviewer. Because we were interested in estimating overdose rates among PWID since 2010, we included only overdose data from person who injected within 12 months of data collection. Exclusion criteria were as follows: [1] study population only includes lifetime (“ever”) non-medical users of injection drugs, [2] findings only include either prevalence or rates of lifetime (“ever”) non-fatal and/or fatal overdose among PWID, [3] study not conducted in an OECD country, [4] study only includes data collected before 2010, [5] study included case reports or overdose descriptions with no denominators, [6] study participants were not human subjects aged 18 years or older, and [7] manuscript not published in English language.

### 2.2 Data Extraction

Data from all eligible studies were extracted into an excel workbook. Data were double extracted from each study, first by one of three researchers (J.Y.S., J.S., M.G.) and then by one of two researchers (E.S. and H.B.). Conflicts were resolved collaboratively. The following data elements were extracted from each study: the number of participants experiencing a non-fatal or fatal overdose; total years of study follow-up; sample size; recall period (non-fatal overdose); recall period (IDU); range/mean/median/IQR for age of participants; the primary objective of the study; whether or not an intervention was evaluated as part of the study; study design; geographic location; study name for participants recruited from an established cohort; date of publication; and first author name.

#### 2.2.1 Quality Assessment

Due to the lack of a standardized quality assessment tool for observational studies, we developed our own tool, adapted from two previous reviews (Bahji et al., 2020; Larney et al., 2020), to evaluate the quality of the data presented in the included studies. The tool assessed each study on the following criteria: whether “overdose” event was clearly defined in the questionnaire (for non-fatal overdose only), how participants were recruited (e.g., population-based vs. venue-based), and whether the number of person-years of follow-up were reported. Evaluation of these study quality domains allowed us to assess the representativeness and accuracy of outcome measures provided by the studies included in the analysis.

### 2.3 Statistical analysis

To have comparable metrics for non-fatal and fatal overdose, we estimated both non-fatal and fatal overdoses in terms of rates per person-years. Non-fatal and fatal overdose rates were calculated by dividing the number of events (i.e., non-fatal or fatal overdoses) by person-years of follow-up in each study. Length of participant follow-up is needed to compute person-years, so for cross-sectional studies, the recall period for non-fatal overdose in the survey instrument was multiplied by sample size to approximate person-years (e.g., 100 people with a 6-month recall period contribute 50 person-years). Reported or approximated person-years were used as denominators for calculating fatal and non-fatal rates, which were log transformed for meta-analysis.

Studies that included overlapping data with other studies meeting inclusion criteria were included in the systematic review but excluded from the meta-analysis; we chose the study with the most inclusive dataset for analysis (see Appendix 6). Notably, thirteen studies that met inclusion criteria reported non-fatal overdose for the same two Canadian cohorts [Vancouver Injection Drug Users Study (VIDUS) and AIDS Care Cohort to Evaluate Exposure to Survival Services (ACCESS)] with varying degrees of temporal overlap. We requested data on non-fatal overdose during 2010 – 2018 from the primary investigators and included those estimates in the meta-analysis in place of the studies with overlapping data.

Pooled rates of non-fatal and fatal overdose among PWID were estimated using a random effects model, given the variation in the included studies’ methods and contexts. Estimates of the variance component were computed using restricted maximum likelihood methods. We also conducted subgroup analyses by region and country using random effects models when data were available from at least five studies. Small study effects were examined using both funnel plots and trim and fill when sufficient data were available. We were unable to examine sensitivity of results to study quality because only two studies and the two estimates from VIDUS and ACCESS cohort data met “satisfactory” criteria (e.g., defined overdose, population-based recruitment, reported person years) for all three quality assessment criteria. Heterogeneity in the incidence rates across studies were assessed using the Q-test for the significance of the variance component, τ^2^, and I^2^. We used R software version 4.0.4 with the metafor (Viechtbauer, 2010) and meta (Balduzzi et al., 2019) packages for the meta-analysis, forest plots, funnel plots and trim and fill.

## Results

The PRISMA flow chart is shown in Figure 1. We screened 13,307 reports, which resulted in 57 studies included in the systematic review, 33 of which contributed unique data to meta-analyses (Figure 1). In total, there were 35 individual estimates in the meta-analysis for non-fatal overdose because we included investigator-contributed VIDUS and ACCESS cohort data to represent the 13 studies with overlapping data on non-fatal overdose from the same cohorts. These 35 estimates represent an approximately equal number of cross-sectional and longitudinal studies, and the majority were from Canada (36%), the U.S. (38%), and Australia (14%) (Table 1). There were considerably fewer estimates available for fatal overdose events (n=8) compared to non-fatal events (n=28).

**Table 1.** Non-Fatal and Fatal Overdose Rates Estimates Among PWID (N=35)

| Author/Publication Year | Country | Study Design | Data Collection Years | N | PY Contributed | Non-Fatal Rate (95% CI) | Fatal Rate (95% CI) |
| --- | --- | --- | --- | --- | --- | --- | --- |
| Betts et al. 2015 | Australia | Cross Sectional | 2011-2013 | 2673 | 222.75 | 47.14 (38.93 — 57.07) |  |
| Horyniak et al. 2013 [a] | Australia | Cross Sectional | 2001-2011 | 6790 | 4707 | 7.90 (7.14 — 8.75) |  |
| Horyniak et al. 2013 [b] | Australia | Cross Sectional | 2008-2010 | 688 | 344 | 19.77 (15.59 — 25.07) |  |
| Iversen et al. 2017 | Australia | Cross Sectional | 2014 | 1488 | 1488 | 20.77 (18.58 — 23.22) |  |
| Kimber et al. 2019 | Australia | Cohort | 1999-2010 | 215 | 107.5; 2095 | 24.19 (16.47 — 35.52) | 0.24 (0.10 — 0.57) |
| Nambiar et al. 2015 | Australia | Cohort | 2008-2012 | 665 | 2277 |  | 0.53 (0.30 — 0.93) |
| Winter et al. 2015 | Australia | Experimental | 2008-2010 | 591 | 292.12 | 27.04 (21.69 — 33.72) |  |
| ACCESS Data | Canada | Cohort | 2010-2018 | 671 | 2341 | 19.18 (17.48 — 21.04) |  |
| Kennedy et al. 2019 | Canada | Cohort | 2006-2017 | 811 | 4928.1 |  | 0.39 (0.25 — 0.60) |
| Selfridge et al. 2019 | Canada | Cohort | 2014-2017 | 132 | 171 |  | 5.26 (2.74 — 10.12) |
| Uhlmann et al. 2014 | Canada | Cohort | 2005-2012 | 1019 | 509.5 | 21.98 (18.26 — 26.45) |  |
| Vallance et al. 2018 | Canada | Cross Sectional | 2008-2015 | 548 | 548 | 18.61 (15.33 — 22.60) |  |
| VIDUS Data | Canada | Cohort | 2010-2018 | 980 | 3434.5 | 24.92 (23.31 — 26.65) |  |
| Wallace et al. 2019 | Canada | Cross Sectional | 2016 | 187 | 93.5 | 59.89 (46.09 — 77.83) |  |
| Rafful et al. 2018 | Mexico | Cohort | 2011-2017 | 671 | 335.5 | 20.03 (15.95 — 25.15) |  |
| West et al. 2020 | Mexico | Cohort | 2011-2018 | 658 | 7471.26 |  | 0.46 (0.33 — 0.64) |
| Folch et al. 2018 | Spain | Cross Sectional | 2014-2015 | 510 | 510 | 19.80 (16.30 — 24.07) |  |
| Åhman et al. 2018 | Sweden | Cohort | 1987-2011 | 2019 | 27698 |  | 0.32 (0.26 — 0.40) |
| O'Halloran et al. 2017 | United Kingdom | Cross Sectional | 2013-2014 | 3850 | 3850 | 15.35 (14.16 — 16.64) |  |
| Trayner et al. 2020 | United Kingdom | Cross Sectional | 2017-2018 | 1437 | 1437 | 18.37 (16.28 — 20.73) |  |
| Al-Tayyib et al. 2017 | United States | Cross Sectional | 2015 | 599 | 599 | 21.37 (17.97 — 25.41) |  |
| Bluthenthal et al. 2020 | United States | Cross Sectional | 2016-2017 | 814 | 407 | 55.53 (48.74 — 63.26) |  |
| Calvo et al. 2017 | United States | Cross Sectional | 2014–2015 | 2251 | 2251 | 7.77 (6.70 — 9.02) |  |
| Carlson et al. 2020 | United States | Cross Sectional | 2017-2018 | 225 | 112.5 | 71.11 (57.12 — 88.53) |  |
| Cedarbaum et al. 2016 | United States | Cross Sectional | 2013 | 389 | 389 | 24.42 (19.97 — 29.86) |  |
| Davis et al. 2017 | United States | Cohort | 1996-2011 | 2007 | 5339 |  | 1.09 (0.84 — 1.41) |
| Hunter et al. 2018 | United States | Cross Sectional | 2016 | 283 | 283 | 32.51 (26.50 — 39.88) |  |
| Latkin et al. 2019 | United States | Cross Sectional | 2016-2018 | 316 | 316 | 35.13 (29.16 — 42.31) |  |
| Oh et al. 2020 | United States | Cross Sectional | 2018 | 458 | 458 | 50.87 (44.74 — 57.84) |  |
| O'Rourke et al. 2019 | United States | Cross Sectional | 2018 | 373 | 186.5 | 87.40 (74.96 — 101.90) |  |
| Riley et al. 2016 | United States | Cohort | 2012-2014 | 173 | 139.6 | 19.34 (13.26 — 28.20) |  |
| Robinson et al. 2017 | United States | Cross Sectional | 2009-2012 | 3457 | 3457 | 13.28 (12.12 — 14.55) |  |
| Rosenthal et al. 2020 | United States | Experimental | 2017-2018 | 100 | 48.33 | 24.83 (14.10 — 43.72) |  |
| Singh et al. 2019 | United States | Cohort | 2001-2015 | 1665 | 13503 |  | 0.47 (0.36 — 0.60) |
| Wagner et al. 2015 | United States | Cross Sectional | 2012-2014 | 573 | 286.5 | 15.71 (11.73 — 21.04) |  |

Person-Years (PY)
Confidence Interval (CI)
\* Non-fatal overdose reported at baseline, fatal overdose measured over follow-up period. Person-years computed from survey look-back for non-fatal but not fatal overdose.
\*\* Excluding unconfirmed deaths
\*\*\* Rates per 100 PY

**Figure 1.**
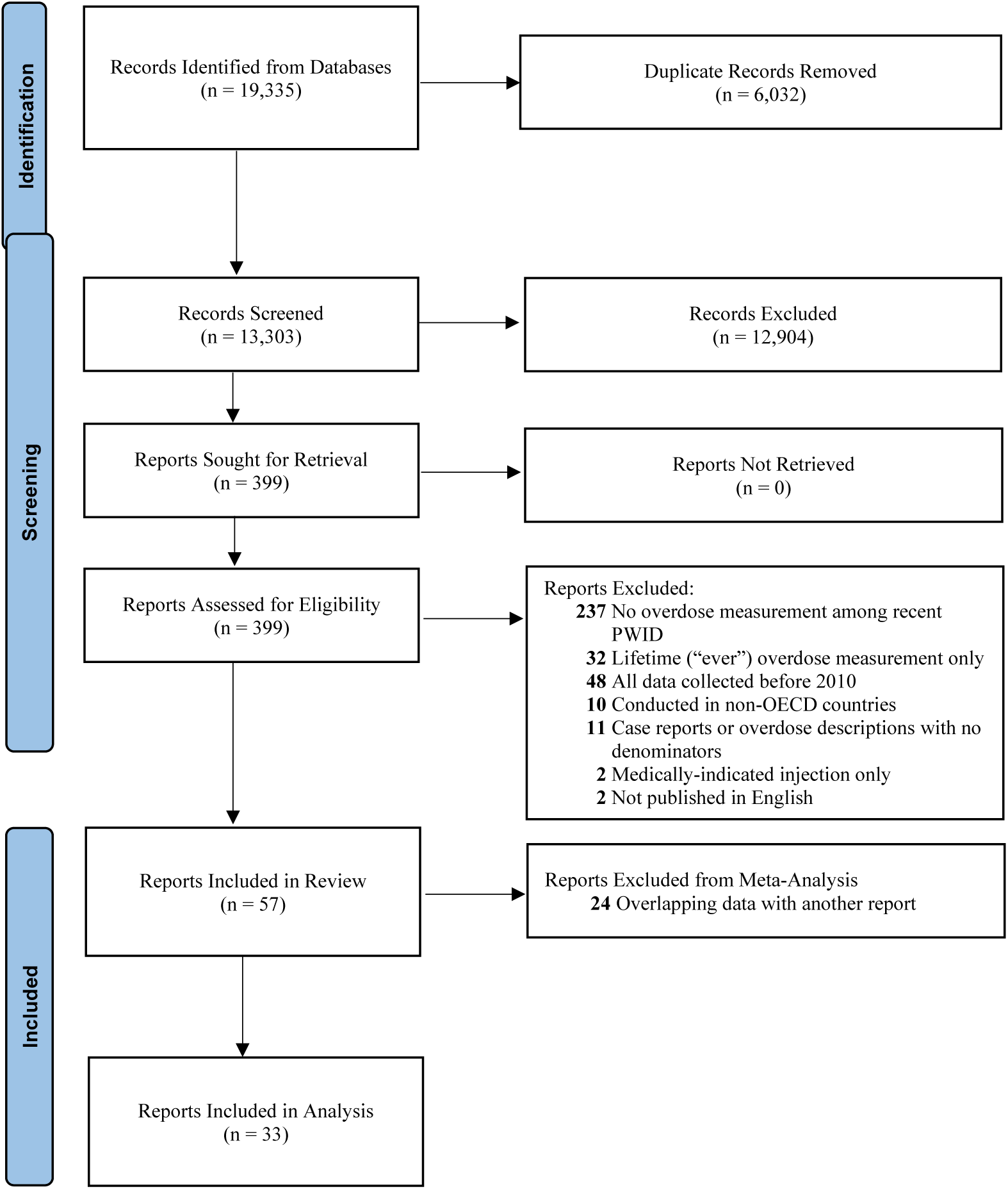
PRISMA Flow Chart.

### 3.1 Non-Fatal Overdose

Twenty-eight estimates contributed to the non-fatal overdose analysis. Across these estimates, there was a mean non-fatal overdose rate of 24.74 per 100 PY (95% CI: 19.86 – 30.83) among PWID living in OECD countries (Table 2). Heterogeneity analysis of the non-fatal rate indicated significant variation across included estimates (Q=1750.91; p-value <0.0001; τ^2^= 0.34). Using I^2^, it was determined that 98.5% of the heterogeniety was due to variability in the effect sizes as opposed to sampling error. The forest plot of these rates (Figure 2) shows individual estimates are similar to the mean, apart from estimates representing a mean data collection year of 2016 or later, in which case many individual rates are above the mean.

**Table 2.** Non-Fatal and Fatal Overdose Rate Estimates (1987-2018)

|  | **n** | **Rate (95% CI)\*** | **$\tau^2$** | **I<sup>2</sup>** | **Q (p-value)** |
| --- | --- | --- | --- | --- | --- |
| **OECD** |  |  |  |  |  |
| Non-Fatal | 28 | 24.74 (19.86—30.83) | 0.34 | 98.5% | 1750.91 (p<0.0001) |
| Fatal | 8 | 0.61 (0.32—1.16) | 0.80 | 93.4% | 106.80 (p<0.0001) |
| **North America** |  |  |  |  |  |
| Non-Fatal | 18 | 27.79 (20.68—37.35) | 0.40 | 98.5% | 1145.45 (p<0.0001) |
| **Australia** |  |  |  |  |  |
| Non-Fatal | 6 | 21.44 (13.32—34.49) | 0.34 | 98.6% | 357.90 (p<0.0001) |
| **Canada** |  |  |  |  |  |
| Non-Fatal | 5 | 25.71 (17.07—38.71) | 0.21 | 94.8% | 76.30 (p<0.0001) |
| **United States** |  |  |  |  |  |
| Non-Fatal | 13 | 28.59 (19.46—42.02) | 0.49 | 98.9% | 1048.51 (p<0.0001) |

\*Estimated from random effects, restricted maximum likelihood models; per 100 PY
n: represents the number of studies contributing data

**Figure 2.**
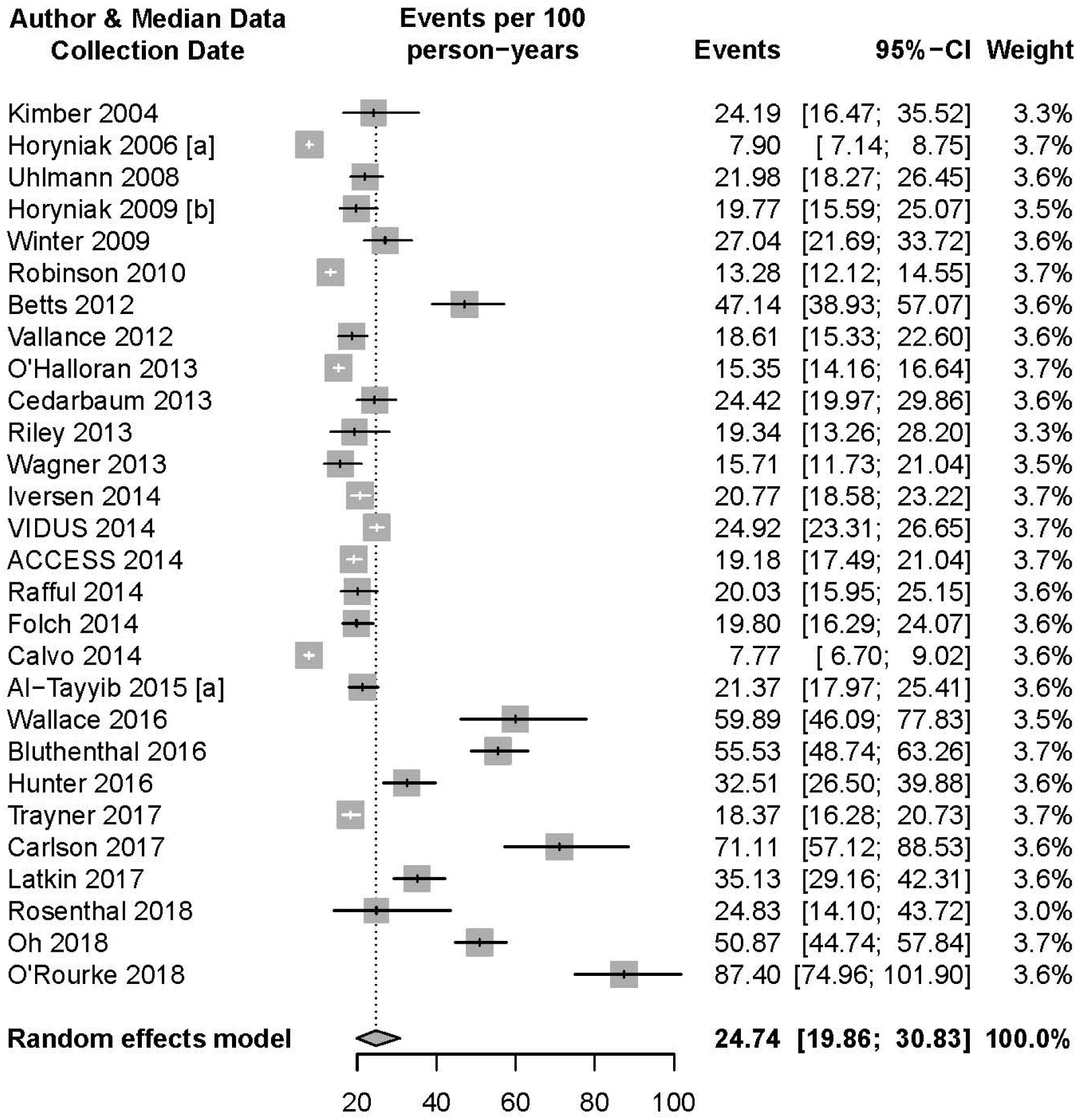
Random-Effects Meta-Analysis of Non-Fatal Overdose Rates Per 100 PY.

Subgroup analysis (Table 1) yielded a mean rate of non-fatal overdose among PWID living in North American countries of 27.79 per 100 PY (95% CI: 20.68 – 37.35). Of the three countries with adequate data to produce country-level estimates, the U.S. had the highest non-fatal overdose rate (28.59 per 100 PY; 95% CI: 19.46 – 42.02), followed by Canada (25.71 per 100 PY; CI: 17.07 – 38.71), and Australia (21.44 per 100 PY; 95% CI: 13.32 – 34.49).

### 3.2 Fatal Overdose

The pooled result, based on 8 studies, showed a mean rate of 0.61 fatal overdoses per 100 PY (95% CI: 0.32 – 1.16) among PWID living in OECD countries (Table 2). As with the non-fatal overdose estimates, the fatal overdose estimates also had significant heterogeneity (Q=106.80; p-value <0.0001; τ^2^= 0.80; I^2^=93.4%) rates (Figure 3). However, the majority of individual study estimates fell close to the mean rate with the exception of one study (Selfridge 2016).

**Figure 3.**
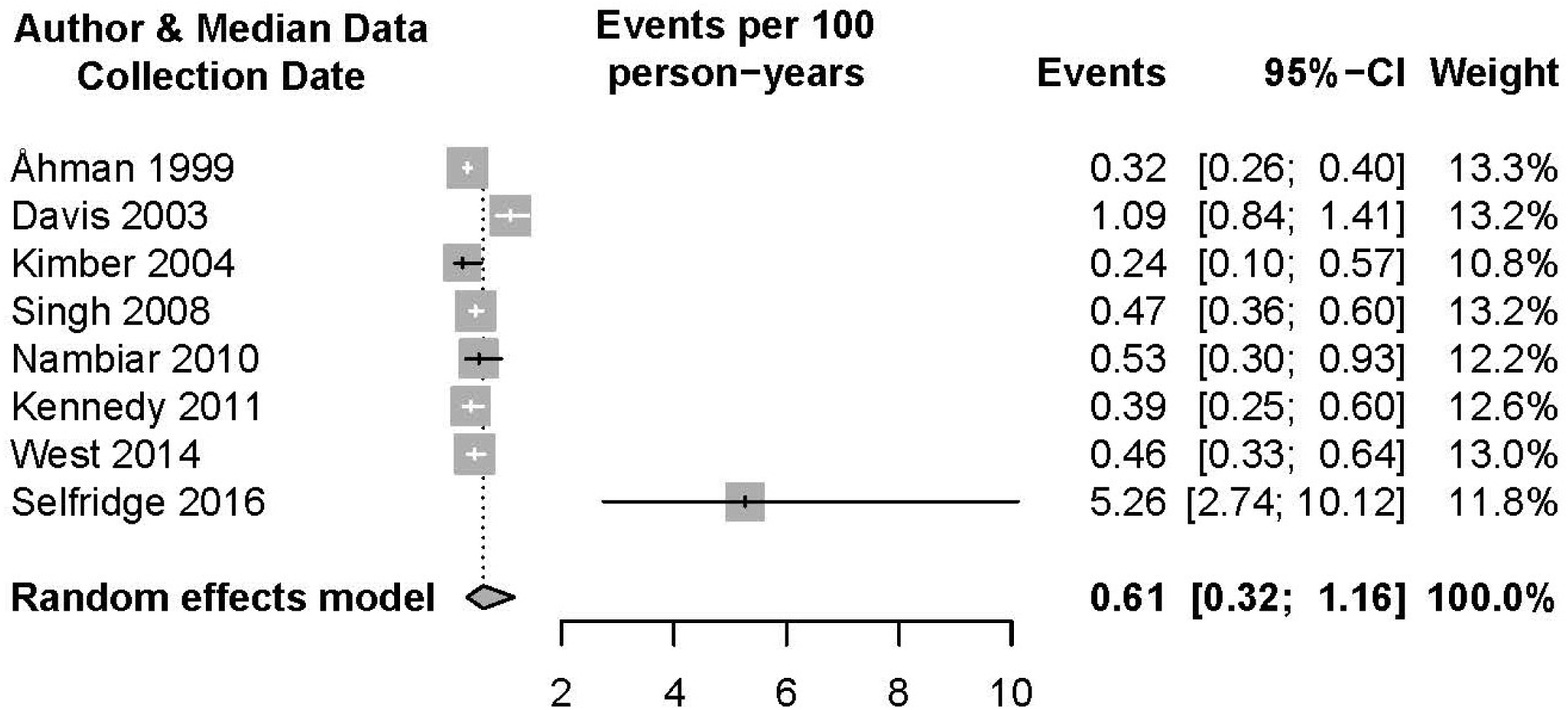
Random-Effects Meta-Analysis of Fatal Overdose Rates Per 100 PY.

### 3.3 Study Quality and Risk of Bias

Over half of the analyzed studies (51%) used population-based recruitment to recruit participants. 34% of studies reported PY contributed by study participants, and 27% of non-fatal overdose estimates were derived from studies that provided participants with a definition of what constituted an overdose event. The trim and fill analysis of the non-fatal overdose estimate included twelve additional effect size estimates, decreasing the mean estimate of the non-fatal overdose to 15.93 per 100 PY (95% CI: 12.15 – 20.89). Given that only eight studies contributed to the fatal overdose estimate, examination of small study effects using a funnel plot was not possible. The trim and fill analysis did not add any effect size estimates to the pooled estimate.

## Discussion

In this systematic review and meta-analysis, we estimated rates of 24.7 non-fatal and 0.6 fatal overdoses per 100 PY among PWID in OECD countries. To our knowledge, this is the first review to estimate rates of both fatal and nonfatal overdose among PWID using comparable methodology. Non-fatal overdose is a strong predictor of fatal overdose (Caudarella et al., 2016), and based on these findings, we estimate one fatal overdose for every 40 non-fatal overdose events among PWID. This ratio provides compelling evidence for the need to prevent not only fatal overdoses, but all overdose events due to the high likelihood of death conditional on overdose (Olfson et al., 2018). This ratio can also be applied to externally derived estimates of either non-fatal or fatal overdose in order to fill data gaps. For example, as data on fatal overdose among PWID become more readily available through surveillance, the ratio can be used to extrapolate the number of fatal overdoses to the corresponding number of non-fatal events (CDC, 2021c; Hall et al., 2021). Additionally, in settings with adequate data on non-fatal overdose burden, the ratio can facilitate evaluation of overdose mortality prevention strategies by allowing assessment of expected versus observed overdose deaths at a given level of non-fatal overdose.

The estimates presented in this review have additional important applications. First, these results can provide comparison estimates to non-fatal and fatal overdose rates among non-injecting substance users, which can be used to understand excess burden of overdose associated with injection. Second, they provide pre-COVID-19 pandemic estimates that can be used as a comparison point for overdose rates among PWID that have likely increased during the pandemic. Quantifying the change in overdose that has occurred during the pandemic is a necessary first step to understanding driving factors (e.g. changes in consumption behaviors, access to harm reduction services) and subsequently intervening on these overdose outcomes (Croxford et al., 2021; Gleason et al., 2021). Third, they can be used to inform resource allocation needs for overdose prevention services such as (SSPs and naloxone distribution, as well as substance use treatment services such as opioid agonist therapy (OAT). Last, these estimates provide a useful baseline estimate for evaluation and effectiveness studies, and implementation models of overdose prevention strategies for PWID.

Harm reduction and substance use treatment services are evidence-based, effective strategies for preventing overdose events among PWID and other people with substance use disorder. The substantial burden of non-fatal and fatal overdose we estimated indicates considerable improvements are needed in provision of harm reduction services, such as SSPs and naloxone distribution, as well as substance use treatment including MAT. Naloxone administration programs have had a notable impact on reducing the burden of overdose among PWID, and a systematic review on take-home naloxone (THN) indicated 95% of naloxone administrations were successful in reversing overdose events (Mcdonald and Strang, 2016). SSPs are also highly effective not only in reducing injection-related infections (by providing sterile injection equipment) but also in increasing naloxone provision and linking PWID to substance use counseling and treatment (Hochstatter et al., 2020; Lambdin et al., 2020). When indicated, OAT can reduce the burden of overdose, as well as other adverse consequences of substance use, among PWID by reducing frequency of substance use (Pearce et al., 2020; Wakeman et al., 2020). Scale-up of overdose prevention and treatment programs, particularly in the U.S., has great potential to save lives, prevent long-term morbidity, and save healthcare costs (Banerjee and Wright, 2020; Saloner et al., 2018).

Very little previous work has been done to estimate non-fatal or fatal overdose rates across recent studies, but there are several relevant comparison points for our estimates. Larney et al. (2020) estimated an all-cause mortality rate of 2.7 per 100 PY among opioid-injecting cohorts globally and a drug-related mortality rate of 0.5 per 100 PY among non-injecting and injecting non-medical users of opioids (Larney et al., 2020). Studies included in this meta-analysis covered a similar time period (2009 – 2019) to our study, but there was no pooled estimate of drug-related mortality among PWID, either globally or by region. Using studies from 2002 – 2017, Colledge et al. (2019) estimated 20.5% of PWID globally experienced at least one overdose in a 12-month period, while 23.0%, 15.6%, and 11.9% experienced at least one overdose per 12 months in North America, Western Europe, and Australia, respectively. Additionally, older systematic reviews and meta-analyses can be used for comparison points over time. Martins et al. (2015) indicated 18.8% of substance users globally, including PWID and others, experienced non-fatal overdose in the past 12 months with fatal overdose rates ranging widely from 0.04 – 46.6 per 100,000 PY (studies published 1980 – 2013) (Martins et al., 2015). Last, Mathers et al. (2013) estimated a global all-cause mortality rate of 0.6 per 100 PY among PWID globally, but this estimate was not presented regionally or by country (Mathers et al., 2013). Although these studies provide important context, simultaneous differences by time, place, and types of substance users (e.g., injecting vs. non-injecting) included make it difficult to directly compare these estimates to our results.

### 4.1 Limitations of The Data

Our systematic review resulted in many studies related to overdose, however studies specific to PWID, or presenting stratified PWID-specific estimates, were limited. Even fewer reported recent overdose among PWID, with many studies including only a lifetime measurement for overdose. These studies were not included because the time periods in which overdose(s) were experienced were unknown. Other studies that focused on PWID were limited to opioid injection, despite recent increases in stimulant injection (Serota et al., 2020). Furthermore, a limitation of studies that did focus broadly on overdose among PWID was the common use of self-report data to measure non-fatal overdose (versus a medically verified event), which may be affected by social desirability bias or an inconsistent understanding of what constitutes an overdose event across participants and interviewers. Resulting information bias would likely lead to an underestimate of non-fatal overdose events.

Most countries included in our review, including the U.S., did not have enough studies to facilitate robust, country-specific meta-analyzed estimates for non-fatal and fatal overdose among PWID. This is problematic because recent data indicate overdose mortality among PWID in the U.S. has increased substantially over time (Hall et al., 2021; Sun et al., 2021). Of the studies included in our review, few included data from the last 2 – 3 years. As a result, we needed to include studies with follow-up periods beginning before recent waves of opioid epidemic (pre-2010) for stable estimates. Even fewer included studies represented rural areas, which have been heavily affected by the U.S. opioid epidemic (Rigg et al., 2018).

### 4.2. Limitations of the Analysis

This review and meta-analysis is also subject to several limitations. First, included studies were limited to peer-reviewed sources, which may have introduced publication bias into our results. However, due to the general dearth of data in this area, we expect most relevant data would be published in the peer-reviewed literature. Second, due to between study variation in risk population and geographic location, we observed a high degree of heterogeneity across study-specific estimates. This heterogeneity may be caused by changes in IDU and substance types over time, as well as country-level differences in response to the epidemic. Last, to produce comparable non-fatal and fatal overdose estimates, we expressed non-fatal overdose as a rate as opposed to a proportion. Where possible, we extracted rates of non-fatal overdose, but many estimates of non-fatal overdose came from cross-sectional survey surveys that asked participants to report any (one or more) overdose in the past year. For these studies, we approximated PY using the recall period and sample size. Doing so may have underestimated non-fatal overdose rate due to an inability to account for multiple overdose events during a given time period.

In recent years, there has been notable investment in the improvement of surveillance data for overdose among PWID in the United States. The primary surveillance source for monitoring non-fatal overdose among PWID is the National HIV Behavioral Surveillance (NHBS) system. NHBS began collecting data on non-fatal overdose in 2018 (CDC, 2020). Additionally, in 2019 CDC invested $12.8 million to improve the timeliness of reporting non-fatal overdose events from emergency medical services and departments (CDC, 2021b). A portion of this investment was allocated to the State Unintentional Drug Overdose Reporting System (SUDORS) to increase the speed and comprehensiveness of reporting fatal overdose events to a national surveillance system (CDC, 2021b). Although these investments improve our ability to monitor unintentional drug overdoses, research studies continue to play an important role in overdose measurement over time. Longstanding cohort studies of PWID are especially helpful for overdose monitoring, but most of these are focused in urban areas. Additional cohort studies across diverse geographic locations, including rural communities, are needed for a more comprehensive understanding of overdose burden. Additionally, mechanisms for harmonizing data elements and sharing data across ongoing cohort studies can help produce more standardized data and shorten the lag time between study completion, publication and use of data to inform timely public health programs.

## Conclusion

In this systematic review and meta-analysis, we report updated estimates of fatal and non-fatal overdose rates among PWID that suggest, in OECD countries similarly affected by the opioid epidemic, there is 1 fatal drug overdose for every 40 non-fatal overdoses. The estimated magnitude of overdose burden reported here underscores the need for expansion of overdose prevention and treatment programs and serves as a baseline estimate for monitoring success of such programs. Given likely increases in IDU and known increases in fatal drug overdose during the COVID-19 pandemic, expansion of surveillance and research on overdose and related health outcomes among PWID should be prioritized.

## Data Availability

All data produced in the present work are contained in the manuscript

## Acknowledgements

The authors acknowledge Marcus Goff and Jonathan Standish for assistance with data extraction. The authors acknowledge funding for this work from the Centers for Disease Control and Prevention, National Center for National Center for HIV, Viral Hepatitis, STD, and TB Prevention (U38PS004650) and the National Institutes of Health, National Institute on Drug Abuse

## Supplemental Materials

## Appendix 1. Search Strings

| Search String | Number of Results |
| --- | --- |
| **PubMed** |  |
| overdose PWID | 165 |
| overdose IDU | 46 |
| fatal* PWID | 53 |
| fatal* IDU | 23 |
| mortal* PWID | 164 |
| mortal* IDU | 177 |
| overdose inject* AND drug | 1134 |
| ((overdose or fatal* or mortal*) AND (inject* or PWID or IDU)) AND (drug* or narcotic) AND (NOT rat or mouse or mice) | 2530 |
| ((((((((((overdose or fatal or mortal) AND (inject or PWID or IDU)) AND (drug or narcotic)))) ) AND ("2010"[Date - Publication] : "3000"[Date - Publication]))) NOT (rat)) NOT (mouse)) NOT (mice) NOT (dog)) NOT (horse) | 1523 |
| **Embase** |  |
| overdose PWID | 204 |
| overdose IDU | 98 |
| fatal* PWID | 56 |
| fatal* IDU | 40 |
| mortal* PWID | 258 |
| mortal* IDU | 310 |
| overdose inject* AND drug | 6262 |
| ((('drug overdose'/exp OR 'drug overdose' OR fatal* OR mortal*:af) AND inject* OR pwid OR 'idu'/exp OR idu) AND drug* OR narcotic) NOT ('rat'/exp OR rat) NOT ('mouse'/exp OR mouse) NOT ('mice'/exp OR mice) NOT ('dog'/exp OR dog) NOT ('horse'/exp OR horse) NOT ('animal'/exp OR animal) AND [1-1-2010]/sd NOT [29-8-2020]/sd | 1631 |
| **PsychInfo** |  |
| overdose PWID | 35 |
| overdose IDU | 14 |
| fatal* PWID | 8 |
| fatal* IDU | 2 |
| mortal* PWID | 15 |
| mortal* IDU | 17 |
| overdose inject* AND drug | 163 |
| ( overdose or poisoning or toxicity ) AND ( fatal or injury ) AND ( mortality or mortality rate or death or death rate ) AND inject* OR ( pwid or idu or injection drug users or injection drug addicts or people who inject drugs ) AND drug use OR narcotics NOT ( case report or case study or clinical case ) NOT rats NOT ( mouse or mice ) | 4164 |

## Appendix 2. Extended Summary Table

| Author/Publication Year | Country | Recruitment Period | End of Survey Follow-Up | PY Contributed | Non-Fatal Events | Fatal Events |
| --- | --- | --- | --- | --- | --- | --- |
| Betts et al. 2015 | Australia | 2011-2013 | 2013 | 222.75 | 105 |  |
| Horyniak et al. 2013 [a] | Australia | 2001-2011 | 2011 | 4707 | 372 |  |
| Horyniak et al. 2013 [b] | Australia | 2008-2010 | 2010 | 344 | 68 |  |
| Iversen et al. 2017 | Australia | 2014 | 2014 | 1488 | 309 |  |
| Kimber et al. 2019 | Australia | 1999-2002 | 2010 | 107.5; 2095 | 26 | 5 |
| Nambiar et al. 2015 | Australia | 2008-2010 | 2012 | 2277 |  | 12 |
| Winter et al. 2015 | Australia | 2008-2010 | 2011 | 292.12 | 79 |  |
| ACCESS Data | Canada | 2006-2014 | 2018 | 2341 | 449 |  |
| Kennedy et al. 2019 | Canada | 2006-2016 | 2017 | 4928.1 |  | 19 |
| Selfridge et al. 2019 | Canada | 2014-2017 | 2017 | 171 |  | 9 |
| Uhlmann et al. 2014 | Canada | 2005-2012 | 2012 | 509.5 | 112 |  |
| Vallance et al. 2018 | Canada | 2008-2015 | 2015 | 548 | 102 |  |
| VIDUS Data | Canada | 2010-2018? | 2018 | 3434.5 |  | 856 |
| Wallace et al. 2019 | Canada | 2016 | 2016 | 93.5 | 56 |  |
| Rafful et al. 2018 | Mexico | 2011-2015 | 2017 | 335.5 | 64 |  |
| West et al. 2020 | Mexico | 2011-2013 | 2018 | 7471.26 |  | 34 |
| Folch et al. 2018 | Spain | 2014-2015 | 2015 | 510 | 101 |  |
| Åhman et al. 2018 | Sweden | 1987-2011 | 2011 | 27698 |  | 89 |
| O'Halloran et al. 2017 | United Kingdom | 2013-2014 | 2014 | 3850 | 591 |  |
| Trayner et al. 2020 | United Kingdom | 2017-2018 | 2018 | 1437 | 264 |  |
| Al-Tayyib et al. 2017 [a] | United States | 2015 | 2015 | 599 | 128 |  |
| Bluthenthal et al. 2020 | United States | 2016-2017 | 2017 | 407 | 226 |  |
| Calvo et al. 2017 | United States | 2014-2015 | 2015 | 2251 | 175 |  |
| Carlson et al. 2020 | United States | 2017-2018 | 2018 | 112.5 | 80 |  |
| Cedarbaum et al. 2016 | United States | 2013 | 2013 | 389 | 95 |  |
| Davis et al. 2017 | United States | 1996-2011 | 2011 | 5339 |  | 58 |
| Hunter et al. 2018 | United States | 2016 | 2016 | 283 | 92 |  |
| Latkin et al. 2019 | United States | 2016-2018 | 2018 | 316 | 111 |  |
| Oh et al. 2020 | United States | 2018 | 2018 | 458 | 233 |  |
| O'Rourke et al. 2019 | United States | 2018 | 2018 | 186.5 | 163 |  |
| Riley et al. 2016 | United States | 2012-2014 | 2014 | 139.6 | 27 |  |
| Robinson et al. 2017 | United States | 2009-2012 | 2012 | 3457 | 459 |  |
| Rosenthal et al. 2020 | United States | 2017-2018 | 2018 | 48.33 | 12 |  |
| Singh et al. 2019 | United States | 2001-2015 | 2015 | 13503 |  | 63 |
| Wagner et al. 2015 | United States | 2012-2014 | 2014 | 286.5 | 45 |  |

Person-Years (PY)
Confidence Interval (CI)

## Appendix 3.1. Quality Assessment Summary (N=35)

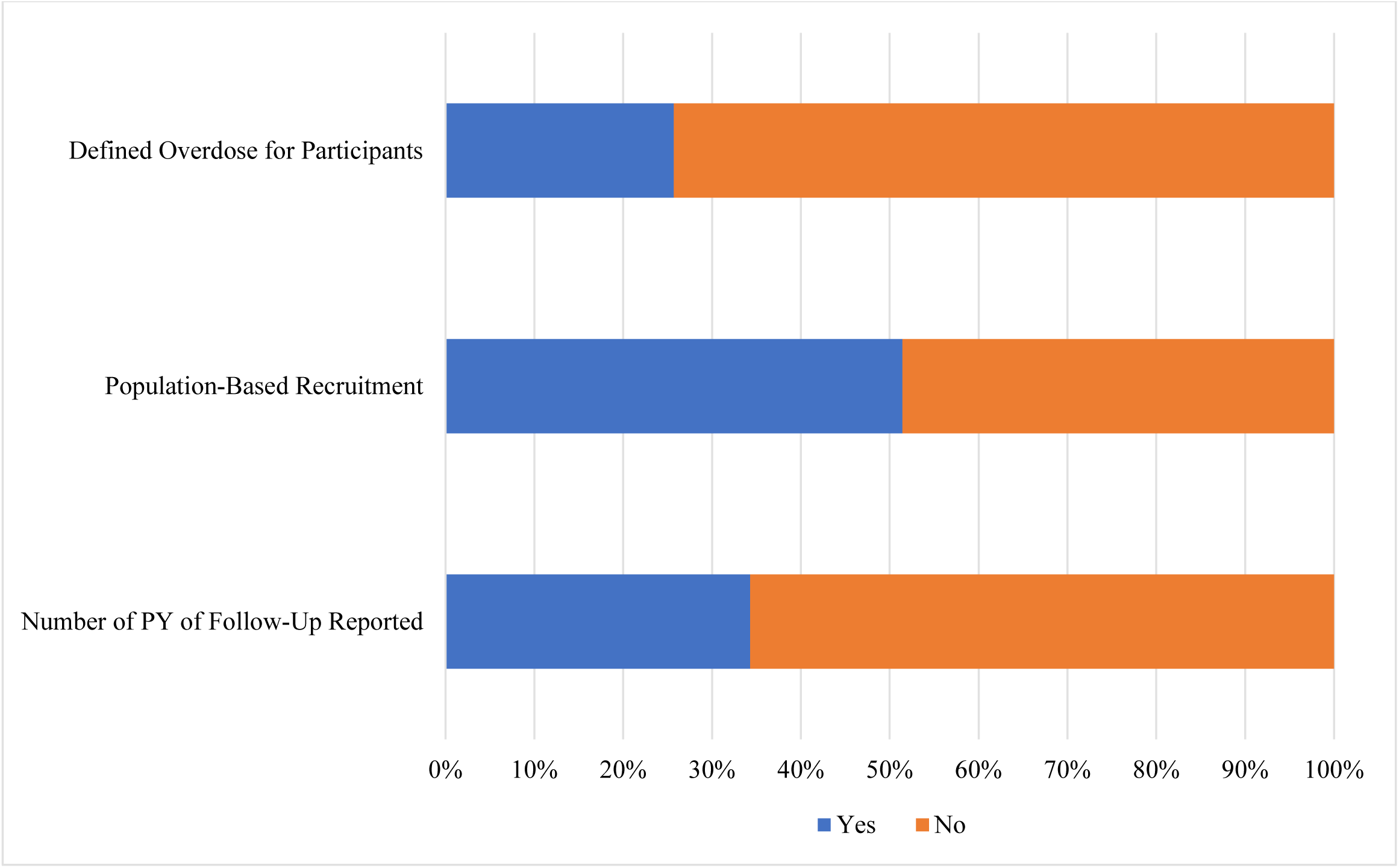

## Appendix 3.2. Non-Fatal Overdose Quality Assessment (N=28)

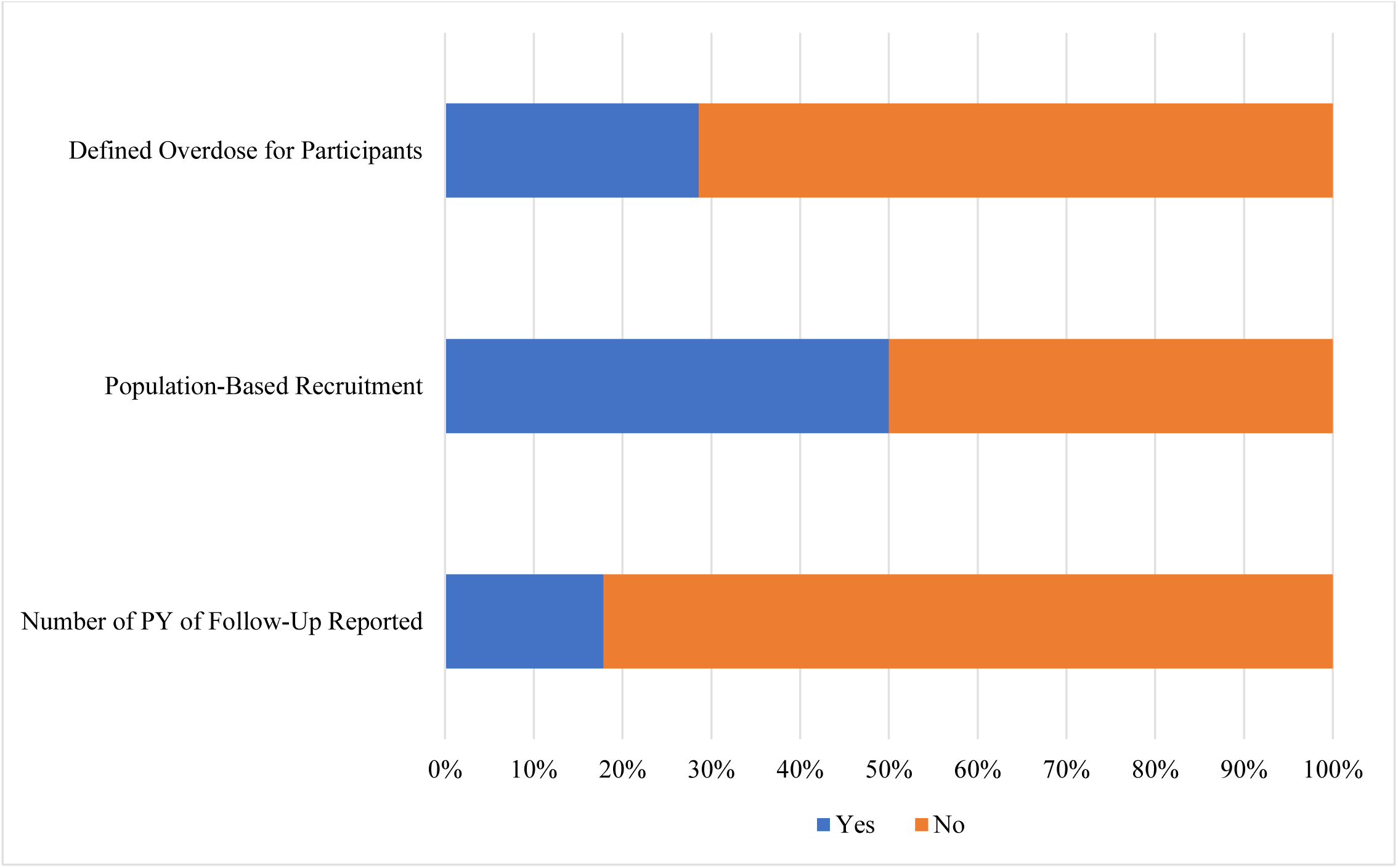

## Appendix 3.3. Fatal Overdose Quality Assessment (N=8)

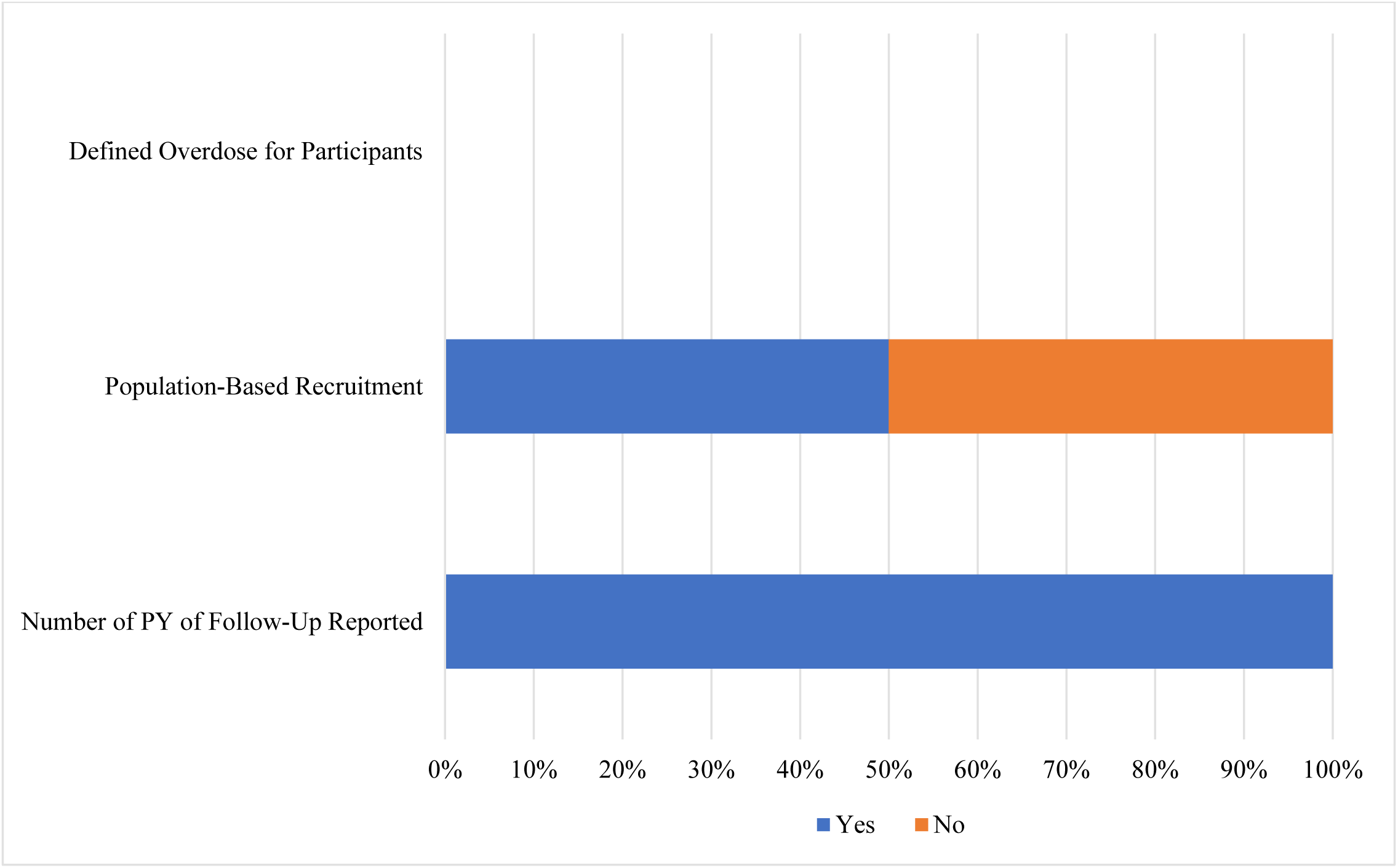

**Table 3.4.**
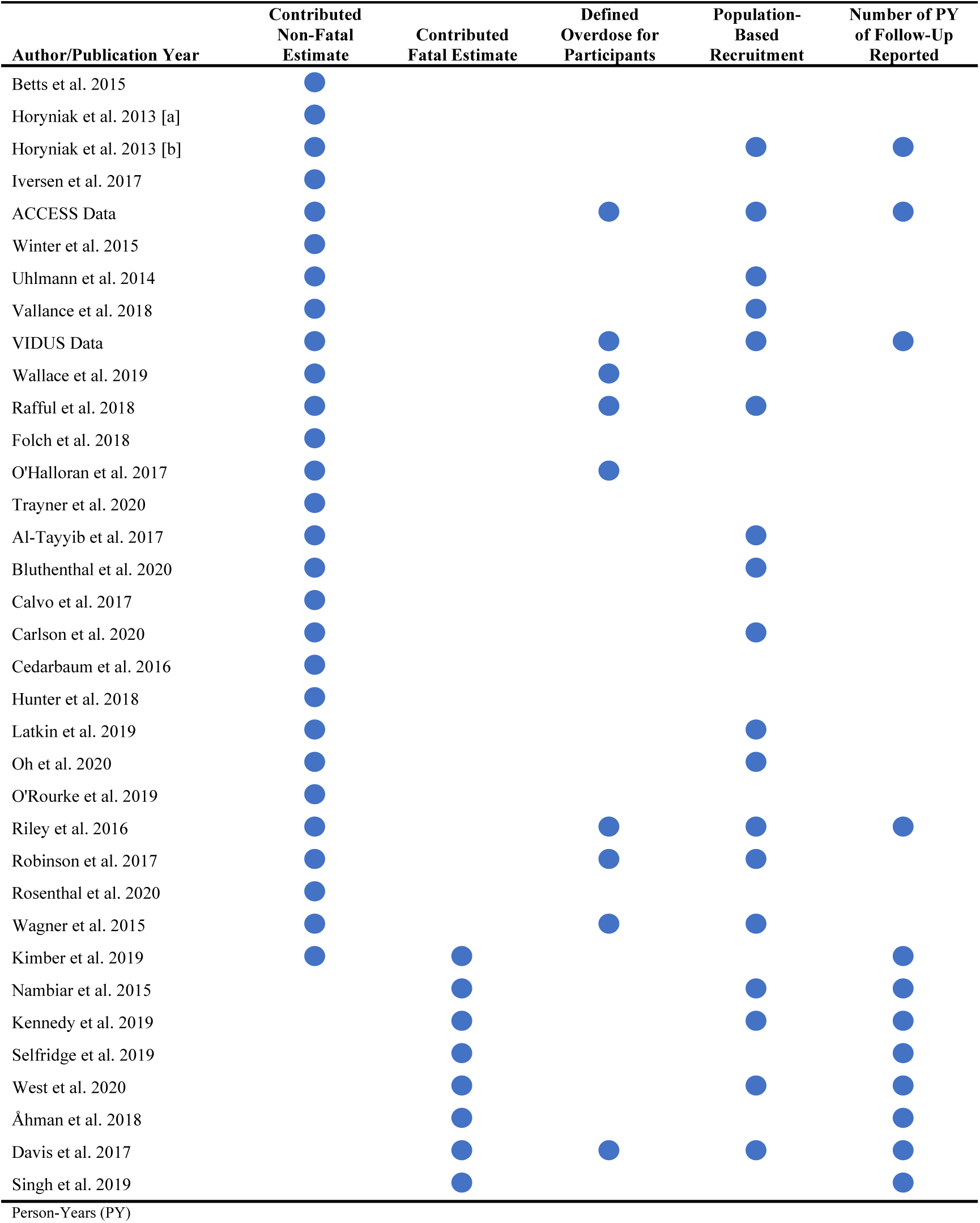
Quality Assessment Scores (N=35)

| Author/Publication Year | Contributed<br>Non-Fatal<br>Estimate | Contributed<br>Fatal Estimate | Defined<br>Overdose for<br>Participants | Population-Based<br>Recruitment | Number of PY<br>of Follow-Up<br>Reported |
| --- | --- | --- | --- | --- | --- |
| Betts et al. 2015 | ● |  |  |  |  |
| Horyniak et al. 2013 [a] | ● |  |  |  |  |
| Horyniak et al. 2013 [b] | ● |  |  | ● | ● |
| Iversen et al. 2017 | ● |  |  |  |  |
| ACCESS Data | ● |  | ● | ● | ● |
| Winter et al. 2015 | ● |  |  |  |  |
| Uhlmann et al. 2014 | ● |  |  | ● |  |
| Vallance et al. 2018 | ● |  |  | ● |  |
| VIDUS Data | ● |  | ● | ● | ● |
| Wallace et al. 2019 | ● |  | ● |  |  |
| Rafful et al. 2018 | ● |  | ● | ● |  |
| Folch et al. 2018 | ● |  |  |  |  |
| O'Halloran et al. 2017 | ● |  | ● |  |  |
| Trayner et al. 2020 | ● |  |  |  |  |
| Al-Tayyib et al. 2017 | ● |  |  | ● |  |
| Bluthenthal et al. 2020 | ● |  |  | ● |  |
| Calvo et al. 2017 | ● |  |  |  |  |
| Carlson et al. 2020 | ● |  |  | ● |  |
| Cedarbaum et al. 2016 | ● |  |  |  |  |
| Hunter et al. 2018 | ● |  |  |  |  |
| Latkin et al. 2019 | ● |  |  | ● |  |
| Oh et al. 2020 | ● |  |  | ● |  |
| O'Rourke et al. 2019 | ● |  |  |  |  |
| Riley et al. 2016 | ● |  | ● | ● | ● |
| Robinson et al. 2017 | ● |  | ● | ● |  |
| Rosenthal et al. 2020 | ● |  |  |  |  |
| Wagner et al. 2015 | ● |  | ● | ● |  |
| Kimber et al. 2019 | ● | ● |  |  | ● |
| Nambiar et al. 2015 |  | ● |  | ● | ● |
| Kennedy et al. 2019 |  | ● |  | ● | ● |
| Selfridge et al. 2019 |  | ● |  |  | ● |
| West et al. 2020 |  | ● |  | ● | ● |
| Åhman et al. 2018 |  | ● |  |  | ● |
| Davis et al. 2017 |  | ● | ● | ● | ● |
| Singh et al. 2019 |  | ● |  |  | ● |
| Person-Years (PY) |  |  |  |  |  |

## Appendix 4. North American Forest Plots

**Figure 4.1.**
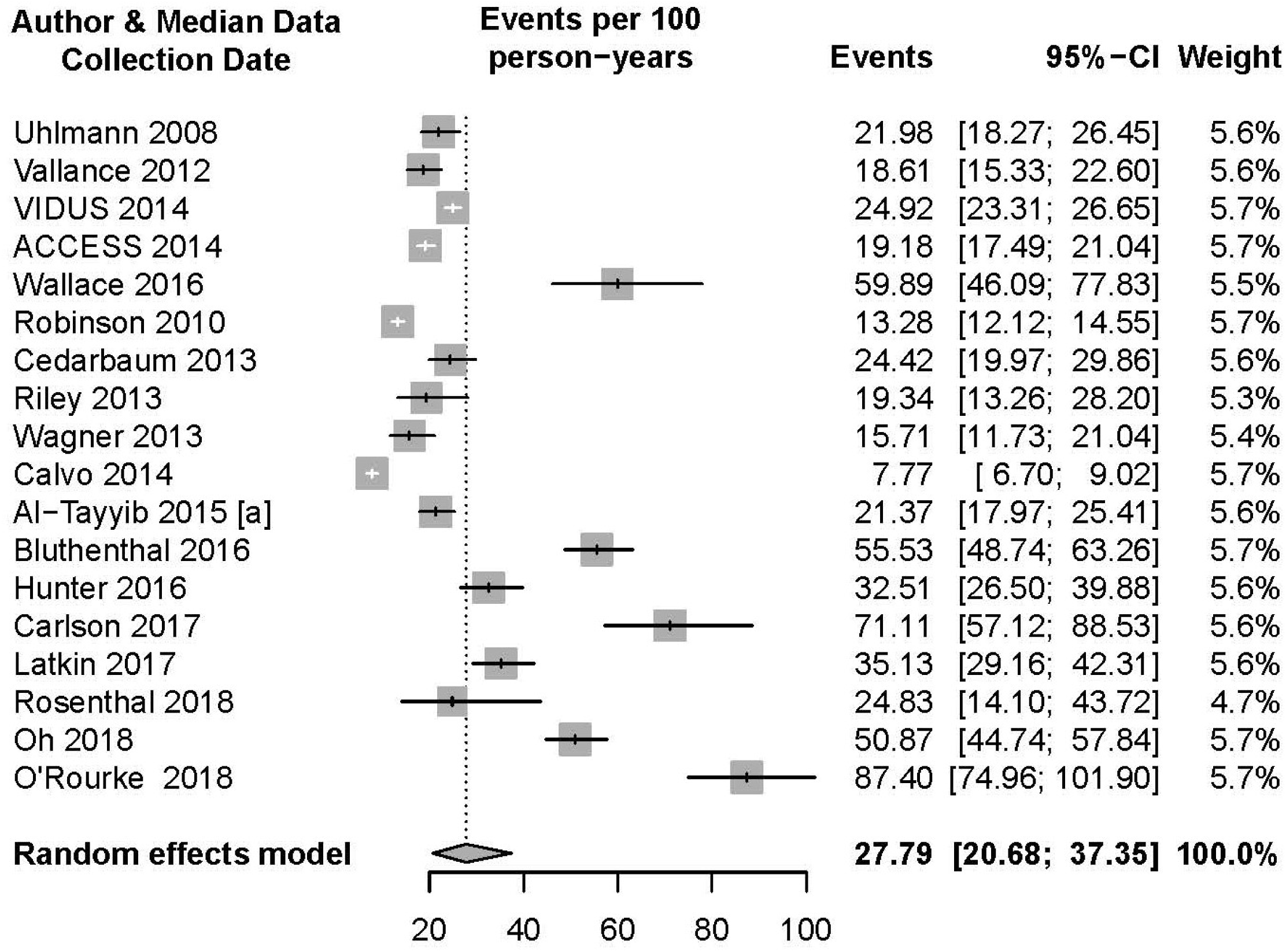
Random-Effects Meta-Analysis of North American Non-Fatal Overdose Rates Per 100 PY

## Appendix 5. Country Specific Forest Plots

**Figure 5.1.**
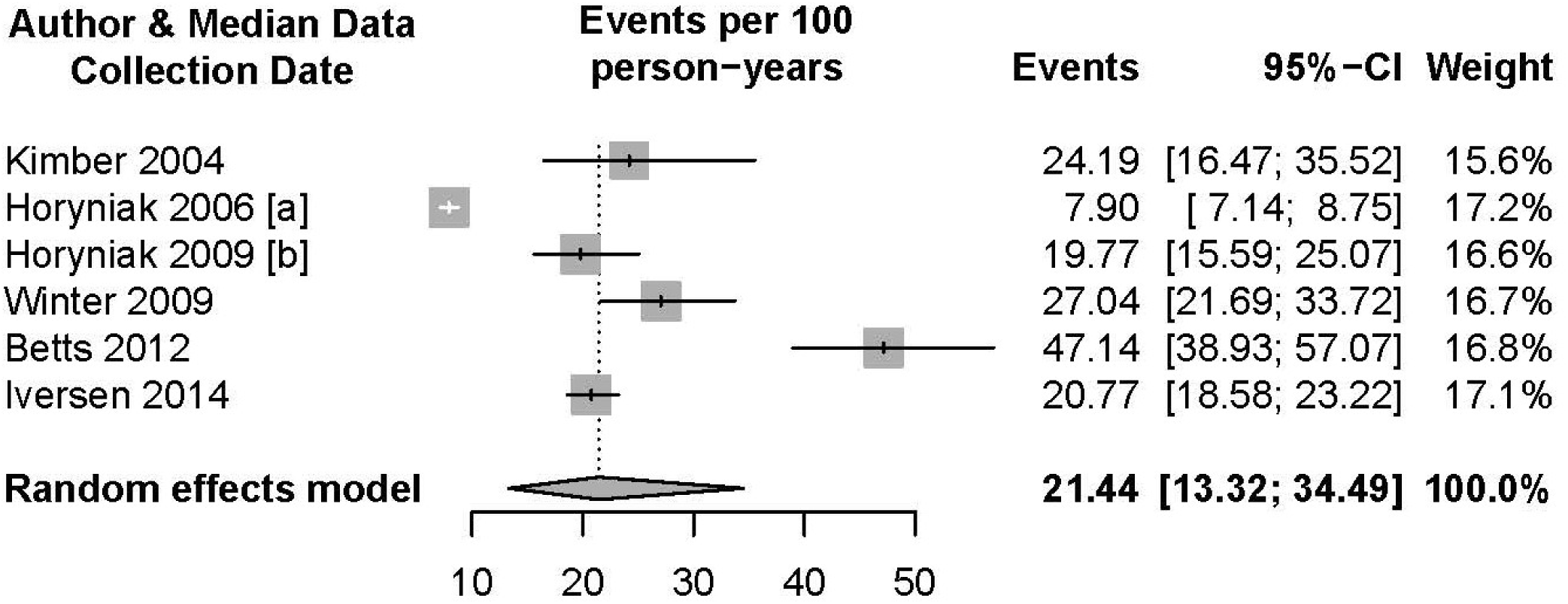
Random-Effects Meta-Analysis of Australian Non-Fatal Overdose Rates Per 100 PY

**Figure 5.2.**
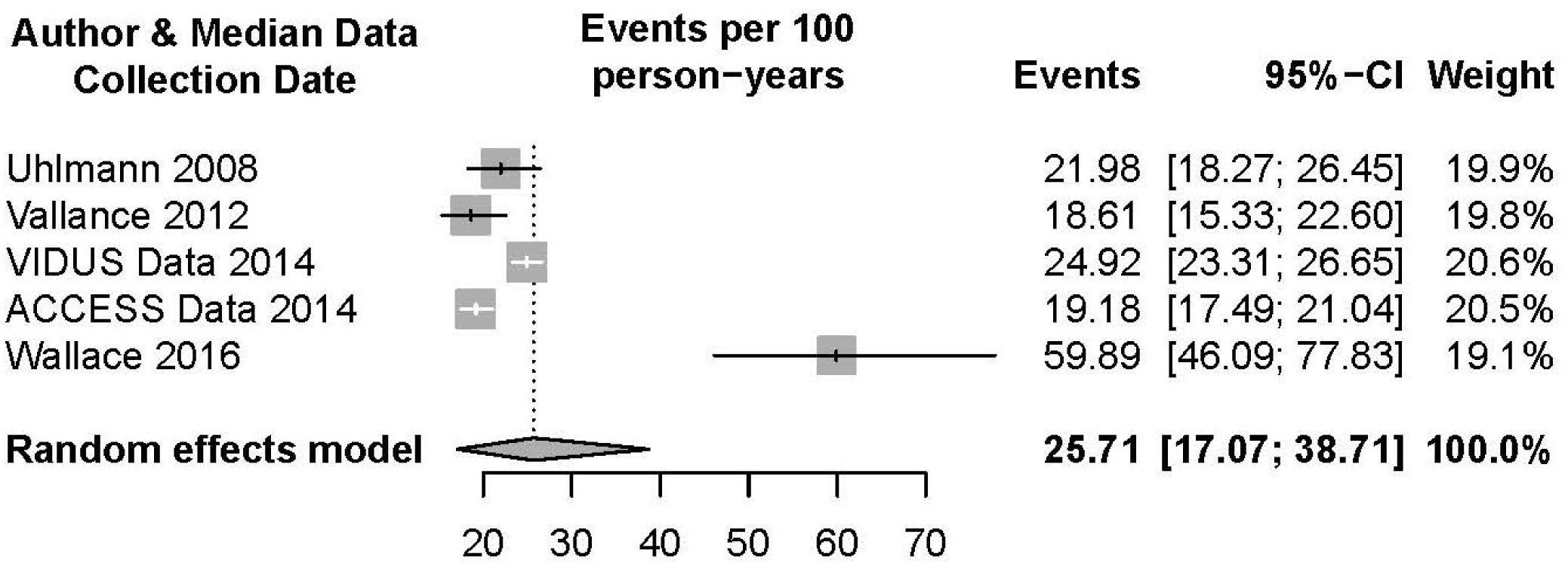
Random-Effects Meta-Analysis of Canadian Non-Fatal Overdose Rates Per 100 PY

**Figure 5.3.**
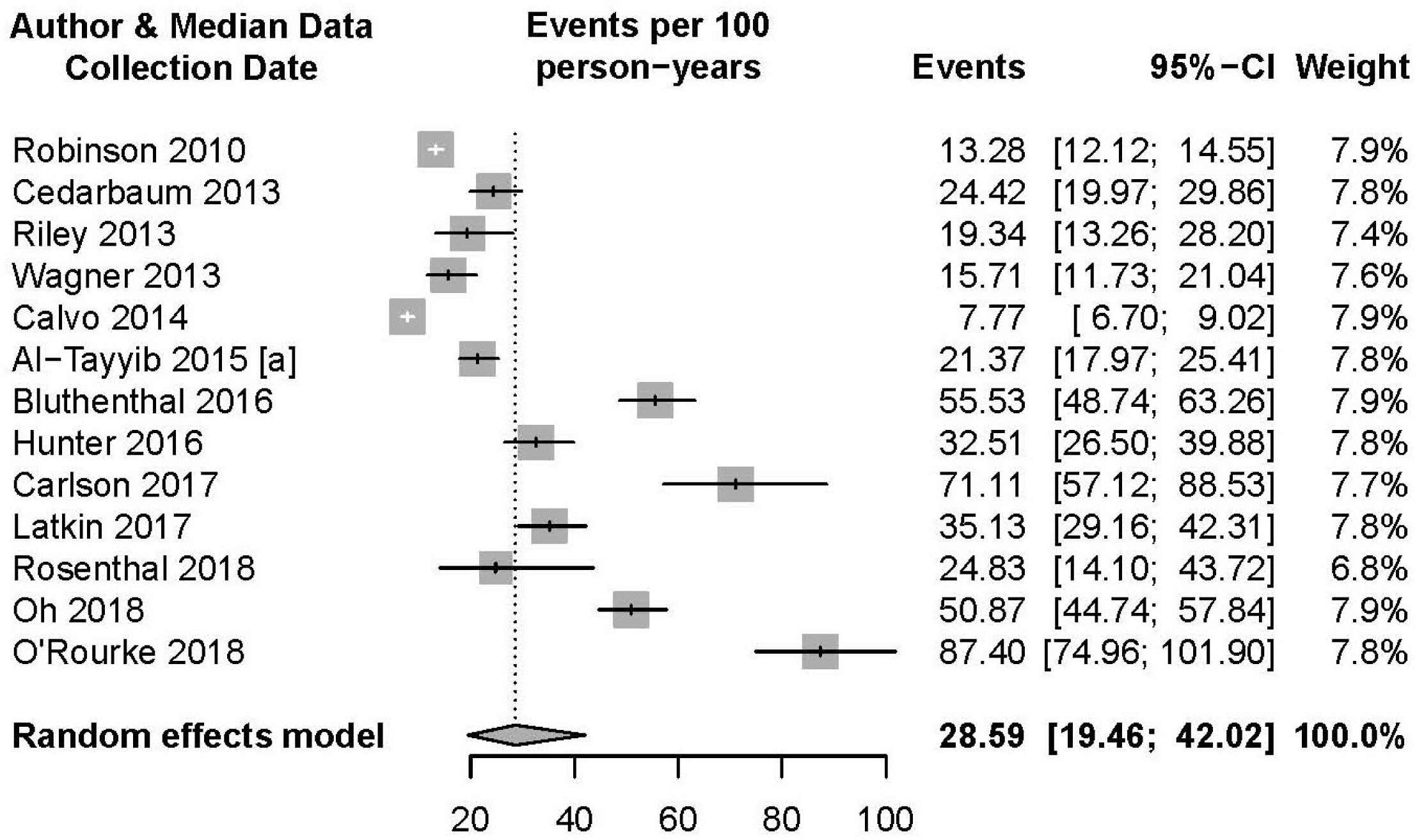
Random-Effects Meta-Analysis of United States Non-Fatal Overdose Rates Per 100 PY

## Appendix 6. Non-Analyzed Data Summary Table (N=24)

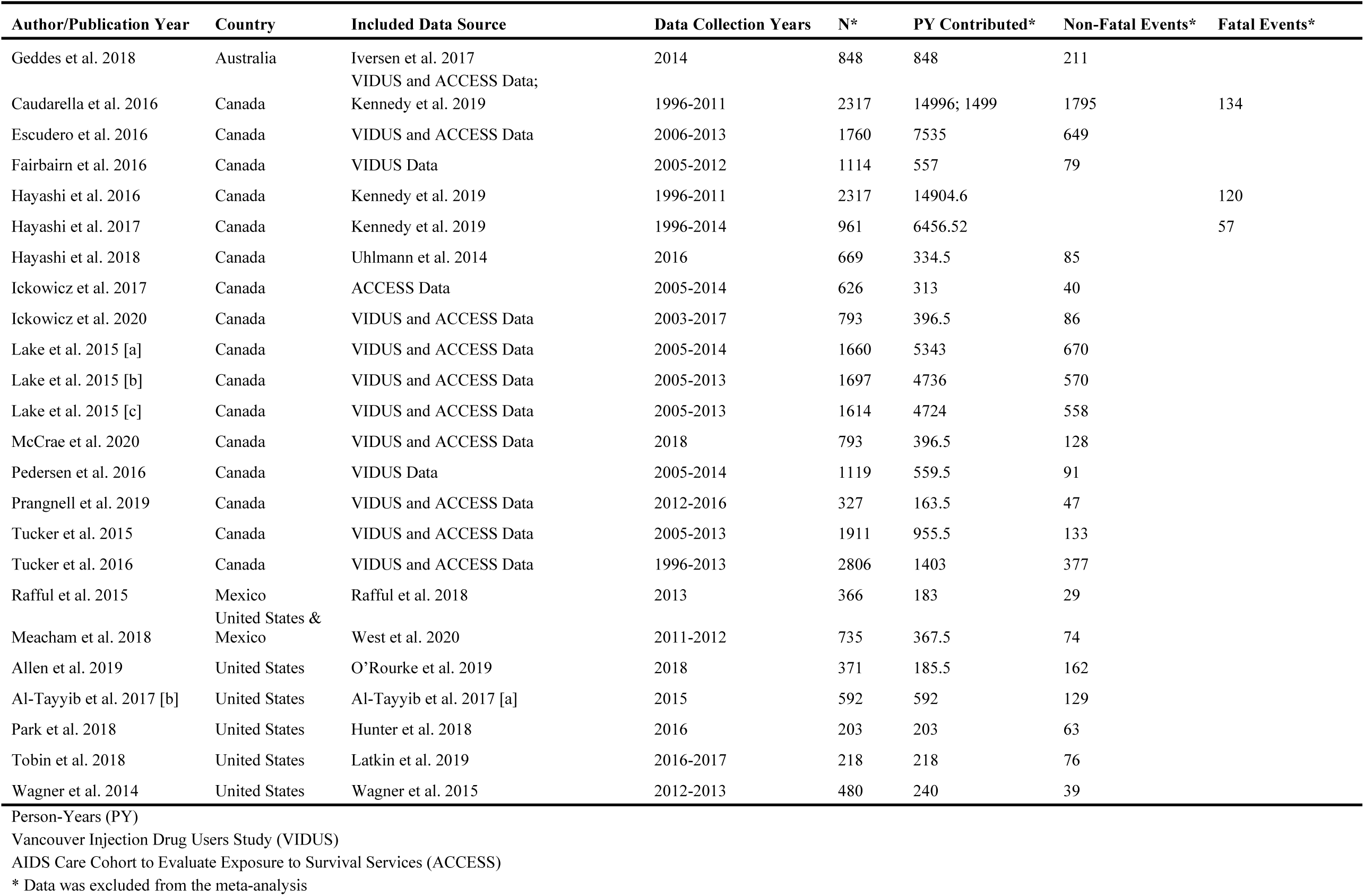

